# Prevalence and determinants of chronic respiratory diseases in adults in Khartoum State, Sudan

**DOI:** 10.1101/2022.06.04.22275986

**Authors:** Rana Ahmed, Nada Osman, Bandar Noory, Rashid Osman, Hana ElHassan, Hind Eltigani, Rebecca Nightingale, Andre FS Amaral, Jaymini Patel, Peter G. Burney, Kevin Mortimer, Asma ElSony

## Abstract

**Background:** Chronic respiratory diseases are considered a significant cause of morbidity and mortality worldwide, although data from Africa are limited. This study aimed to determine the prevalence and determinants of chronic respiratory diseases in Khartoum, Sudan.

**Methods:** Data was collected from 516 participants, aged _≥_ 40, who had completed a questionnaire and undertook pre- and post-bronchodilator spirometry testing. Trained field workers conducted questionnaires and spirometry. Survey-weighted prevalence of respiratory symptoms and spirometric abnormalities were estimated. Regression analysis models were used to identify risk factors for chronic lung diseases.

**Results:** Using the NHANESIII reference equations, the prevalence of Chronic Airflow Obstruction (CAO) was 10%. The main risk factor was older age 60-69 years (Odds ratio 3.16, 95% Confidence Interval 1.20 – 8.31). Lower education, high body mass index and a history of tuberculosis were also identified as significant risk factors. The prevalence of a low forced vital capacity (FVC) using NHANES III was 62.7% [SE 2.2] and 11.3% [SE 1.4] using locally derived values.

**Conclusion:** The prevalence of spirometric abnormality mainly (low FVC); was high suggesting that chronic respiratory disease is of substantial public health importance in urban Sudan. Strategies for the prevention and control of these problems are needed.

---

The Global Burden of Disease Study estimates 3.9 million deaths annually from chronic non-communicable respiratory diseases – mainly chronic obstructive pulmonary disease (COPD) and asthma^1,2^. It’s burden of deaths and morbidity is expected to increase over future decades especially in low-income and middle-income countries (LMICs)^3,4^.

Prevalence estimates for COPD in sub-Saharan Africa (SSA)^5,6^ are based on limited epidemiological data which lack a standardized definition of COPD. A recent systematic review reported a population prevalence of COPD in SSA ranging from 1.7% to 24.8%^7^.

The prevalence and determinants of low FVC in SSA are barely understood however, its reported that Africans have reduced FVC compared with the Caucasian^8^. Moreover, studies reported an association between lung restriction and mortality and a higher prevalence of Chronic Respiratory Disease (CRD) in SSA that linked to numerous risk factors including early childhood exposures, poverty, biomass fuel exposure, smoking and pulmonary tuberculosis (TB) ^9^.

The Burden of Obstructive Lung Disease (BOLD) Initiative developed standardized methods for estimating the burden and determinants of chronic airflow obstruction (CAO) in populations aged 40 years and older^10,11^. We did a BOLD study in Khartoum, Sudan to help fill the knowledge gap about the chronic respiratory diseases in Africa.

## METHODS

### Setting

Sudan’s capital, Khartoum, is made up of 7 localities, across which there is a mix of urban, semi-urban, rural, and internally displaced populations.

### Sampling

Three localities, with a total population of 661,617, were randomly selected for sampling in this study and divided into clusters. Then 280, 258 and 158 households were randomly selected from Jabelawlya, Shargalneel and Omdurman localities respectively.

### Participants

Using the BOLD protocol^11^, we approached 998 participants from which 600 _≥_ 40 years old were included using a 3-stage stratified cluster sampling plan. Potential participants who were institutionalized or medically unfit to perform spirometry were excluded.

### Data collection and management

All study participants completed a structured interview in the local Arabic language administered by trained interviewers. Anthropometric measurements along with pre-bronchodilator and post-bronchodilator spirometry data were collected following the American Thoracic Society (ATS) guidelines using the Easy One system (ndd Medizintechnik, Zurich, Switzerland) by three trained certified technicians^11^. A minimal data or refusal questionnaire was filled out for those not willing to participate in the full study. The clinical data obtained included height, weight, pulse rate and waist and hip circumference. Quality control was carried out at the BOLD coordinating centre. Usable spirometry was defined as two or more acceptable blows, with FEV_1_ and FVC repeatability within 200 mL. Acceptable manoeuvres were defined as those with a rapid start (back-extrapolated volume, 150 mL or 5% of the FVC), lack of cough during the first second, and a small end-of-test volume (<40 mL during the final second). The calibration of all spirometers was verified to be accurate within 3.0% using a 3.00 L syringe at the beginning of each day of testing. Spirometry traces were then classified according to FEV_1_/FVC < lower limit of normality (LLN).

CAO was defined by post-bronchodilator (BD) FEV_1_/FVC < LLN. Predicted values based on standardized values for age, sex, and height were calculated based on the Third National Health and Nutrition Examination Survey 1988– 1994 (NHANES III) of white Americans^12^. Local values were derived from spirometry of non-smoking Sudanese adults with no respiratory symptoms or diagnoses participating in this survey. CAO stages were categorised as: stage 1 or higher CAO (Post-BD FEV_1_/FVC < LLN) and stage 2 or higher CAO (Post-BD FEV_1_/FVC < LLN and post-BD FEV_1_ < 80% predicted).

### Statistical analysis

Evaluation of selection bias and a comparison between groups was conducted using a chi-square test between participants who completed full data and minimal questionnaires with acceptable or unacceptable spirometry readings. Prevalence estimates of spirometric abnormalities stratified by age and sex were reported using the NHANES III ^11^. Prevalence estimates using locally derived spirometry were also reported.

Univariable and multivariable logistic regression analyses were used to test associations between spirometry abnormalities and several exposure variables, including age, sex, education level, self-reported history of tuberculosis (TB), hypertension, diabetes, heart disease, body mass index, smoking status, smoking pack-years, exposure to indoor smoke from biomass fuel and occupational exposure.

A wealth score was developed based on the Mokken scale, to differentiate between different levels of wealth using a count of owned assets^13,14^.

Multivariable logistic regression models that included sex, age and all variables from the univariable analysis with a *p*-value <0.2 were then developed. The prevalence of respiratory symptoms was reported and associations with the study variables were tested using regression analysis. A description of the associations between abnormal spirometry and respiratory symptoms was reported. The data were analysed using Stata IC 14 (StataCorp, College Station, TX). Prevalence estimates and regression models were developed using survey weighting with the Svy package in Stata (14).

### Ethical considerations

Written informed consent was collected from study participants before data collection. Ethical approval was obtained from the Imperial College London and Khartoum state Ministry of Health.

## RESULTS

The flow of participants through the study is shown in Figure 1. Of the 998 participants approached, 516 provided full questionnaire data and had BOLD centre-approved spirometry results. Eleven of the 998 participants declined to participate fully in the study but completed the minimal data questionnaire. The final response rate was 85.5% (n=696).

**Figure 1.** Participant flow diagram

### Participant characteristics

These are presented in Table 1. The mean age was 53.8 years (SD 10.4) and 59.3% were men. Overall, 35% completed primary school. Men had a higher level of education, as did the group aged 40-49 years when compared to other age groups. The mean number of household members was 7.8 (SD3.56) and the mean wealth score was 5.2 (SD 2.7).

**Table 1.**
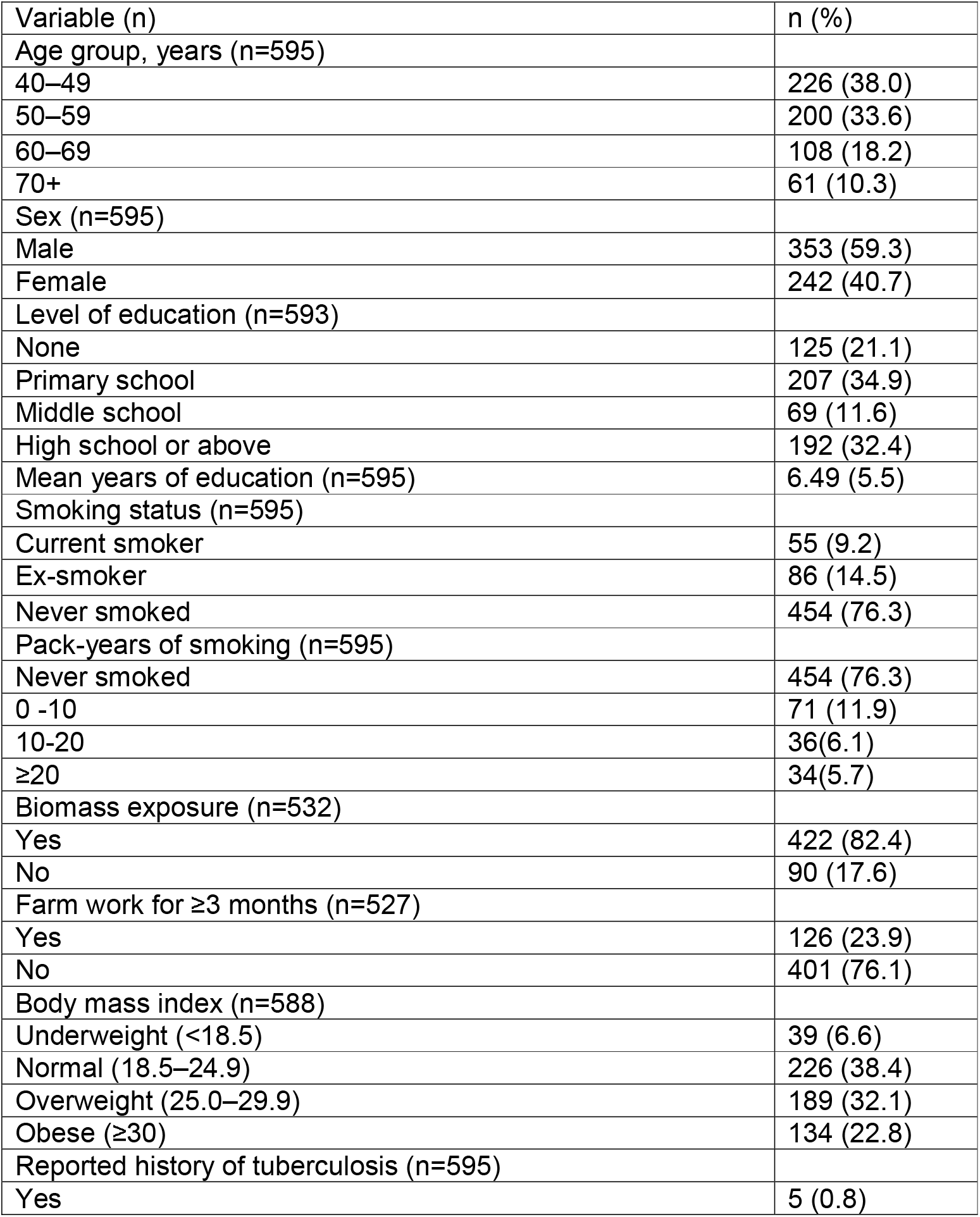
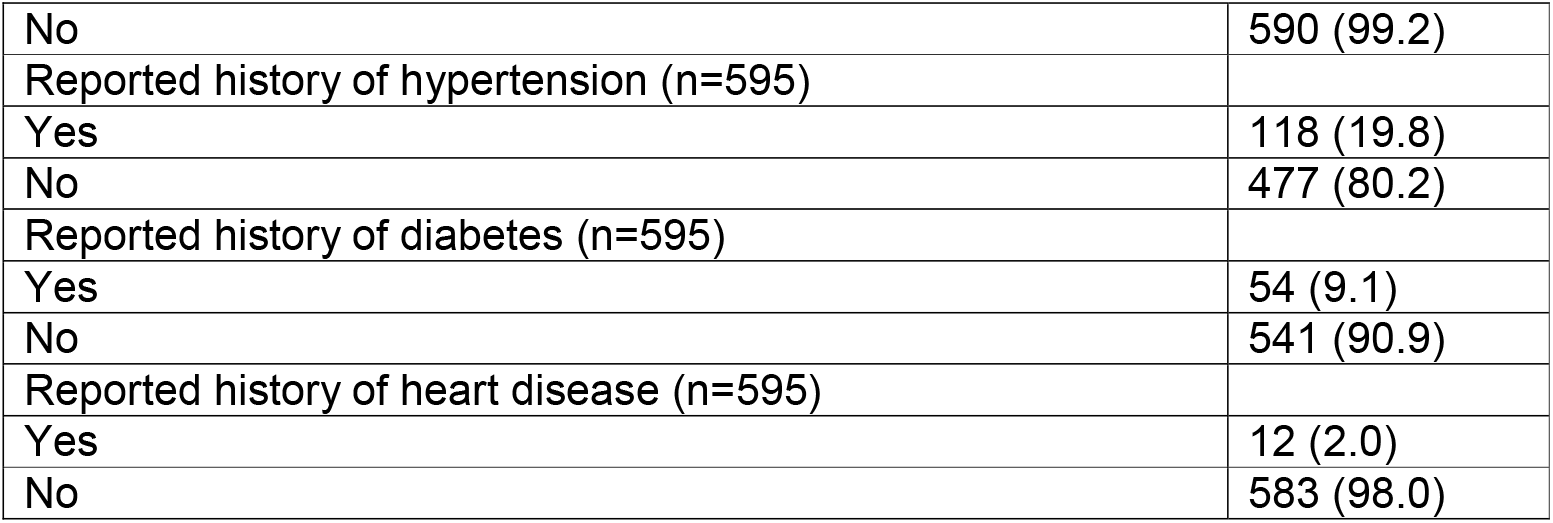
Characteristics of all subjects who completed a full BOLD core questionnaire, including those with and without spirometry results

| Variable (n) | n (%) |
| --- | --- |
| Age group, years (n=595) |  |
| 40–49 | 226 (38.0) |
| 50–59 | 200 (33.6) |
| 60–69 | 108 (18.2) |
| 70+ | 61 (10.3) |
| Sex (n=595) |  |
| Male | 353 (59.3) |
| Female | 242 (40.7) |
| Level of education (n=593) |  |
| None | 125 (21.1) |
| Primary school | 207 (34.9) |
| Middle school | 69 (11.6) |
| High school or above | 192 (32.4) |
| Mean years of education (n=595) | 6.49 (5.5) |
| Smoking status (n=595) |  |
| Current smoker | 55 (9.2) |
| Ex-smoker | 86 (14.5) |
| Never smoked | 454 (76.3) |
| Pack-years of smoking (n=595) |  |
| Never smoked | 454 (76.3) |
| 0 -10 | 71 (11.9) |
| 10-20 | 36(6.1) |
| ≥20 | 34(5.7) |
| Biomass exposure (n=532) |  |
| Yes | 422 (82.4) |
| No | 90 (17.6) |
| Farm work for ≥3 months (n=527) |  |
| Yes | 126 (23.9) |
| No | 401 (76.1) |
| Body mass index (n=588) |  |
| Underweight (<18.5) | 39 (6.6) |
| Normal (18.5–24.9) | 226 (38.4) |
| Overweight (25.0–29.9) | 189 (32.1) |
| Obese (≥30) | 134 (22.8) |
| Reported history of tuberculosis (n=595) |  |
| Yes | 5 (0.8) |
| No | 590 (99.2) |
| Reported history of hypertension (n=595) |  |
| Yes | 118 (19.8) |
| No | 477 (80.2) |
| Reported history of diabetes (n=595) |  |
| Yes | 54 (9.1) |
| No | 541 (90.9) |
| Reported history of heart disease (n=595) |  |
| Yes | 12 (2.0) |
| No | 583 (98.0) |

Among respondents, 24% had smoked cigarettes while 50% of men were current smokers. About 24% of smokers had more than a 20 pack-years of exposure. Exposure to indoor biomass fuel for _≥_ 6 months was reported by 82% of participants. In total, 78% reported having used an indoor open fire fuelled by coal or charcoal for cooking for _≥_ 6 months, while 35% used firewood and 21% used kerosene. Overall, women had a higher mean number of hours of exposure to indoor biomass fuel per year than men (70% vs. 54%). Farming was the most reported occupation (in 24%, Table 1).

In total, 23% of participants were obese and 7% were underweight. Hypertension was self-reported by 20% of all participants, of whom 55% were women. Diabetes was reported by 9% (9.5% of women and 8.8% of men).

### Respiratory symptoms

At least one respiratory symptom was reported by 23% (Standard error (SE) 1.9) of participants; respiratory symptoms interfering with daily activities were reported by 1.9% (SE 0.5). Cough was reported by 10.4% (SE 1.3), with the highest prevalence recorded in participants aged 70+ years (11.9% [SE 4.9]). Chronic cough (lasting for more than 3 months per year) was reported by 4.0% (SE 0.8). Production of sputum was reported by 11% (SE 1.3) and chronic production of sputum (for more than 3 months per year) was reported by 5% (SE 0.09). Shortness of breath was reported by 11% (SE 1.3) and 41% (SE 6.5) of this group reported that breathing problems made it difficult to walk more than 100 yards. Wheeze in the past 12 months in the absence of cold was the least commonly reported symptom (3.0% [SE 0.7], Supplementary Table S1).

### Spirometry

No statistically significant differences were found between the groups who did or did not complete the spirometry test. Using NHANES III, stage 1 or higher CAO prevalence was 10.3% [SE 1.4] (9.2 [SE 1.7] of men and 11.2 [SE 2.4] of women). Using the locally derived reference range the prevalence was 5.7% [SE 1.1] (5.2% [SE 1.3] of men and 6.2 [SE 1.9] of women).

Participants aged 60-69 years had the highest prevalence of stage 1 or higher CAO (13.4% [SE 3.8]). Prevalence of stage 2 or higher CAO was 9.4% [SE 1.4] (8.8% [SE 1.7] of men and 10.1% [SE 2.2] of women). Using the locally derived reference range, 3.0% [SE 0.8] of the study population had stage 2 or higher CAO (2.9% [SE 0.9] of men and 3.1% [SE 1.3] of women). Similarly, participants aged 60-69 years had the highest prevalence of stage 2 or higher CAO using both the local and NHANES reference ranges (17.6% [SE 4.2] vs. 6.7% [SE 2.6]).

Low FVC was seen in 62.7% [SE 2.2] (65.2% [SE 2.8] in men vs. 59.8% [SE 3.5] in women). Cough was less reported in those with low FVC (OR 0.48, 95% CI 0.27 - 0.87).

Airflow reversibility was found in 6.1% [SE 1.1] of the total study population and was more common in women than men (8% [SE 1.9] vs. 4.4% [SE 1.1]). Airflow obstruction persisted after use of a bronchodilator in 8.4% (SE 4.7) of participants with reversibility. (Supplementary Table S2, Figure 2).

**Figure 2.** Estimated Population Prevalence of chronic airflow obstruction by age and sex using National Health and Nutrition Examination Survey reference ranges (NHANES) for the Sudanese population in participants completing standard American Thoracic Society spirometry (n=516). The upper graph represents the prevalence of Stage 1 or higher CAO (Post-BD FEV1/FVC < LLN) and the lower graph represents the prevalence of Stage 2 or higher CAO (Post-BD FEV1/FVC < LLN and post-BD FEV1 < 80% predicted).

### Factors associated with respiratory symptoms

In both univariable and multivariable analyses, chronic production of sputum was negatively associated with age (Supplementary Tables S3 and S4). Participants aged 60-69 years were less likely to report chronic sputum production (OR 0.39, 95% CI 0.16 - 0.93) than those aged 40-49 years. There was a significantly increased likelihood of regular sputum production with being an ex-smoker (OR 2.66, 95% CI 1.09 - 6.50) and having diabetes (OR 4.04, 95% CI 1.82 - 8.96). Participants with lower socioeconomic status; who have a wealth score of 2 tend to have higher odds of sputum production compared to those with zero score (OR 7.18, 95% CI 1.16 - 44.53).

In multivariable analysis, the likelihood of having shortness of breath was significantly greater in participants exposed to indoor biomass fuel (OR 4.56, 95% CI 1.44 - 14.43). The presence of wheeze was only associated with being a current smoker (OR 3.49, 95% CI 1.02 - 11.96). There were no significant associations between CAO and respiratory symptoms.

### Factors associated with post-bronchodilator airway obstruction

Participants aged 60-69 years had the highest risks of CAO stage 1 or higher (OR 3.16, 95% CI 1.20 - 8.31) and stage 2 or higher (OR 3.39, 95% CI 1.04 -n 6.93) than those aged 40-49 years. In contrast, having higher education level was protective against any obstruction in bivariate analysis (OR 0.31, 95% CI 0.13 - 0.76), however no association was identified after adjustment. Similarly, being overweight or obese was protective against any obstruction (OR 0.38, 95% CI 0.17 - 0.82 and OR 0.34, 95% CI 0.13 - 0.99, respectively). Those with a history of TB were less likely to have any obstruction (OR 0.08, 95% CI 0.01 - 0.59). There was no observed trend in Mokken scale points and CAO. However, participants with a low socioeconomic position who scored ≥2 in wealth score had the highest odds of developing stage 1 or higher and stage 2 or higher CAO (OR 6.00, 95% CI 1.03 - 34.94). No other factor was significantly associated with airway obstruction (Table 2, Supplementary Tables S5 and S6).

**Table 2.** Multivariable associations of risk factors with stage 1 or higher CAO defined using NHANES III (Post-BD FEV_1_/FVC < LLN; n=53/516) and Stage 2 or higher CAO defined using NHANES III (Post-BD FEV1/FVC < LLN and post-BD FEV_1_ < 80% predicted; n=49/516)

| Variable | Multivariable association<br>with CAO stage 1 or<br>higher |  | Multivariable association<br>with CAO stage 2 or<br>higher |  |
| --- | --- | --- | --- | --- |
|  | OR | 95% CI | OR | 95% CI |
| Age group, years |  |  |  |  |
| 40–49 | 1.0 | - | 1.0 | - |
| 50–59 | 2.13 | 0.84 - 5.41 | 1.86 | 0.73 - 4.77 |
| 60–69 | 3.16 * | 1.20 - 8.32 | 2.78* | 1.07 - 7.26 |
| 70+ | 1.91 | 0.60 - 6.10 | 0.91 | 0.21 - 3.92 |
| Sex |  |  |  |  |
| Male | 1.0 | - | 1.0 | - |
| Female | 1.31 | 0.61 - 2.84 | 1.67 | 0.76 - 3.66 |
| Level of education |  |  |  |  |
| None | 1.0 | - | 1.0 | - |
| Primary school | 0.61 | 0.27 - 1.34 | 0.62 | 0.27 - 1.44 |
| Middle school | 1.23 | 0.40 - 3.81 | 1.03 | 0.32 - 3.36 |
| High school or above | 0.71 | 0.28 - 1.78 | 0.69 | 0.26 - 1.86 |
| Body mass index |  |  |  |  |
| Underweight (<18.5) | 1.87 | 0.66 - 5.30 | 1.16 | 0.33 - 4.07 |
| Normal (18.5–24.9) | 1.0 | - | - | - |
| Overweight (25.0–29.9) | 0.43 | 0.18 - 1.01 | 0.37* | 0.15 - 0.92 |
| Obese (≥30) | 0.35 | 0.11 - 1.17 | 0.29* | 0.08 - 1.07 |
| Self-reported TB |  |  |  |  |
| No | 1.0 | - | 1.0 | 0 |
| Yes | 0.08* | 0.01 - 0.59 | 0.07* | 0.01 - 0.48 |
| Number of people living in | ± | ± | 1.05 | 0.96 - 1.16 |
| Used firewood |  |  |  |  |
| No | ± | ± | 1.0 | - |
| Yes | ± | ± | 0.58 | 0.29 - 1.19 |
| Wealth score/Mokken scale | ± | ± | 0.96 | 0.83 - 1.11 |

\* $p < 0.05$ . CAO, Chronic Airflow Obstruction; CI, confidence interval; OR, odds ratio; FEV<sub>1</sub>, forced expiratory volume in 1 second; FVC, forced vital capacity ratio; LLN, lower limit of normal, ( ± ) not included in multivariable analysis

Using the local reference range, participants aged 60-69 years were more likely to have CAO stage 1 or higher than their younger counterparts (OR 3.10, 95% CI 1.01 - 9.57), and being obese were negatively associated with obstruction (OR 0.29, 95% CI 0.09 - 0.97) in multivariable analysis. A higher education level was protective against CAO in the bivariate analysis (OR 0.23, 95% CI 0.063 - 0.83) however no association was identified after adjustment.

### Factors associated with low FVC

Using the NHANES III, low FVC was associated with smoking 10-20 packs year history, having primary or higher-level education, having more people in house and being obese (OR 2.79, 95% CI 1.11 - 7.00; OR 2.42, 95% CI 1.43 -4.09; OR 0.94, 95% CI 0.89 - 0.99 and OR 1.73, 95% CI 1.04 - 2.86 respectively) in bivariate analysis. In multivariate analysis, those aged 50-59 were less likely to have low FVC compared to those aged 40-49 years (OR 0.50, 95%CI 0.31 - 0.81). No other factors were associated with low FVC in multivariate analysis (Table 3).

**Table 3.** Multivariable associations of risk factors with low FVC, defined using NHANES III reference range (FVC < LLN), n= 315/516

| Variable | Multivariable association |  |
| --- | --- | --- |
|  | OR | 95% CI |
| Age group |  |  |
| 40-49 | 1 | - |
| 50-59 | 0.50* | 0.31 - 0.81 |
| 60-69 | 0.29* | 0.17 - 0.51 |
| 70+ | 0.25* | 0.12 - 0.53 |
| Sex |  |  |
| Male | 1 | - |
| Female | 0.78 | 0.48 - 1.24 |
| Body Mass Index (kg/m<sup>2</sup>) |  |  |
| Underweight (BMI < 18.5) | 0.95 | 0.43 - 2.07 |
| Normal (BMI 18-25) | 1 | - |
| Overweight (BMI 25-30) | 1.39 | 0.85 - 2.27 |
| Obese (BMI > 30) | 1.55 | 0.87 - 2.80 |
| Packs-year of smoking |  |  |
| Never | 1 | - |
| 0-10 | 1.09 | 0.59 - 2.02 |
| 10-20 | 2.45 | 0.88 - 6.86 |
| $\geq 20$ | 1.00 | 0.47 - 2.11 |
| Wealth score/Mokken scale | 0.97 | 0.90 - 1.04 |

\* $p < 0.05$ . CI, confidence interval; OR, odds ratio; FVC, forced vital capacity ratio; LLN, lower limit of normal

## DISCUSSION

This population-based survey aimed to investigate the prevalence and determinants of chronic respiratory diseases in the population aged _≥_40 years in urban Sudan. Our main finding was that approximately 10% of people in this age group had CAO. CAO stage II or higher was detected in 9.4% of the overall population using the same reference range, though this decreased to 3.0% when the local reference range was used.

A higher prevalence of obstruction when using the NHANES III compared to the local reference range has been reported previously^9^. In spite of being the only available data, values of the local methodology might be more ethnically suitable compared to NHANES III. However, different exposures in this setting may constrain its use^9,15^.

The determined prevalence of spirometric obstruction is consistent with that in similar studies from sub-Saharan Africa, where the prevalence of smoking is high^16^. Previous BOLD studies reported COPD prevalence of 23% in men and 16.9% in women in South Africa^17^ and 7.7% in Nigeria^7,18,19^. CAO prevalence reported from a multinational BOLD study was 11.5% in men and 8.8% in women which is consistent with our findings^20^. Compared to studies from the MENA region (Middle East and North Africa), our findings are higher than those in Saudi Arabia^21^, Tunisia^22,23^, Morocco^24^, Algeria^25^ and Lebanon^26^.

Older age was the main risk factor for CAO in our study, which is consistent with both regional and global findings^7,9,16,18^. The absence of an association with CAO and cigarette smoking might be due to that 50% of the smokers in our study reported a smoking history of _≤_10 pack-years. Countries with lower smoking rates, such as Malawi and Rwanda, have a lower reported prevalence of COPD^9,27^. However, the age group 60-69 years who reported the highest smoking rates in this study had the highest prevalence of spirometric obstruction and higher prevalence of both chronic cough and chronic phlegm. The high prevalence of CAO might be due to exposure to air pollutants coming from the large number of factories and cars in the State.

A higher educational level was protective against CAO which is compatible with other studies^10,18,28^. Conversely, a significant association between low socioeconomic status and developing CAO was found in this study, consistent with studies suggesting that low socioeconomic status may be associated with a progression of airflow limitation^16,29^.

We did not observe any association between exposure to biomass fuel and obstruction. This is consistent with the study that included 25 BOLD sites which found that airflow obstruction measured by post-bronchodilator spirometry was not associated with use of solid fuels for cooking or heating^30^, and findings in other large studies in China^31,32^. A recent review of studies on COPD and household air pollution concluded that it was not possible to define clear causal links between the two^33^.

Shortness of breath, one of the most common symptoms of CAO, was significantly higher in those who use biomass fuel for _≥_ 6 months. It is possible that this latter group suffer from chronic bronchitis or similar non-obstructive lung diseases. In addition, the finding that only 0.2% reported TB might account for the significant negative association between TB and obstruction, which is in conflict with published literature^34^. Identification of TB was based on self-reporting and many factors might affect the validity of the answers provided given that TB is a highly stigmatized clinical condition and the proportion reported here may be an underestimate^35^.

A high prevalence of low FVC (62.7%) was identified in this study. BOLD studies in SSA and other studies in resource-poor settings reports similar findings ^9,17^. That 55% of the study population was overweight might partly explain the high level of low FVC^8^.

To our knowledge, this is the first study to provide a detailed prevalence estimates of CAO in Sudanese adults using internationally standardised methods and procedures as well as an appropriate sampling technique. We acknowledge our study had limitations. Missing information for a proportion of the study participants limited the cluster-weighted analysis. Furthermore, reasons for exclusion were not recorded, meaning those who were excluded for medical reasons were not separated from those who were excluded for other reasons. Having spirometry readings with no matching questionnaire data for 54 participants and unacceptable spirometry readings from 79 participants decreased the sample size and study power.

In conclusion, this study found evidence that chronic respiratory disease is a major problem in Sudan and needs to be considered in future public health policies and research. The overall prevalence of CAO in urban Sudan is similar to that found by other BOLD studies in countries with similar smoking rates. However, it is relatively high when compared to other countries in sub-Saharan Africa and the MENA regions. A high prevalence of low FVC was also identified, the aetiology and pathophysiology of which are unknown and require further investigation. These findings suggest the need for strengthening the chronic respiratory disease programs and provision of improved diagnostic and treatment options for CRDs to address underestimation and diagnosis.

## Data Availability

All data produced in the present study are available upon reasonable request to the authors.

## ACKNOWLEDGEMENTS

This work was supported by Welcome Trust, UK - grant (085790/Z/08/Z) for the

BOLD (Burden of Obstructive Lung Disease) study. Prof. Mortimer received advisory board fees from AstraZeneca outside this work and Dr Nightingale reports grants from Medical Research Council, outside the submitted work.

We thank the teams of data collectors and spirometry technicians from Sudan for their hard work during the household survey. We also thank the members of the BOLD Coordinating Centre at the Imperial College London for its assistance with spirometry training, quality control, and data management in the study.

## CONTRIBUTION STATEMENT

Dr Rana Ahmed wrote the first draft of manuscript, conducted data cleaning, verification, interpretation and analysis of this study.

Dr Nada Osman lead the study and contributed to manuscript writing and revision.

Dr Bandar Noory coordinated the study, supervised the data collection and contributed to manuscript writing and revision.

Dr Rashid Osman revised the spirometry readings, contributed to the writing and revision.

Ms Hind Eltigani contributed to data collection, data verification and participated in writing.

Ms Hana ElHassan was responsible for the field administration of the overall project. She contributed to the writing and revision.

Dr Rebecca Nightingale contributed to the writing and analysis of this manuscript.

Dr Andre FS Amaral reviewed and contributed to the writing of this manuscript.

Mrs. Jaymini Patel provided the clean data, analysis report and revised the manuscript.

Prof. Burney reviewed and contributed to the methodology of the study and reviewed the writing of this manuscript.

Prof. Kevin Mortimer supervised, reviewed and contributed to the writing of this manuscript.

Prof. Asma El Sony was the principal investigator of the study, she supervised, reviewed and contributed to the writing of this manuscript.

