## Supplementary table for "Prevalence and determinants of chronic respiratory diseases in adults in Khartoum State, Sudan"

**Table S1**: Age and gender stratified prevalence of respiratory symptoms among participants with complete core questionnaire

| Definition of restriction | Age group | Male(n=353) Prevalence (SE) | Female (n=242) Prevalence (SE) | Total(n=595) Prevalence (SE) |
| --- | --- | --- | --- | --- |
| Cough (Do you usually cough when you don't have a cold?) | 40-49 | 9% (2.7) | 9.1% (2.6) | 9.4% (1.9) |
|  | 50-59 | 10% (2.7) | 10.6% (3.4) | 10.7% (1.9) |
|  | 60-69 | 12% (3.8) | 9.9% (5.4) | 11.6% (3.2) |
|  | 70+ | 11% (4.7) | 13.1% (8.8) | 11.9% (4.9) |
|  | Total | 10.6% (1.6) | 10.1% (2.0) | 10.4% (1.3) |
| Sputum (Do you usually bring up phlegm from your chest?) | 40-49 | 16% (3.4) | 10.8% (2.8) | 13.6% (2.3) |
|  | 50-59 | 10.9% (2.7) | 5.8% (2.5) | 8.6% (1.9) |
|  | 60-69 | 10.2% (3.2) | 13% (6.1) | 11.4% (3.2) |
|  | 70+ | 11.9% (5.1) | 0 | 6.4% (2.9) |
|  | Total | 13.2% (1.9) | 8.5% (1.7) | 11.1% (1.3) |
| Wheeze (Have you had wheezing / whistling in your chest at any point in past 12m, in the absence of a cold) | 40-49 | 4.5% (2.0) | 3.3% (1.6) | 4% ( 1.3) |
|  | 50-59 | 1.7% (1.1) | 1.1%(1.1) | 1.4 (0.8) |
|  | 60-69 | 7.6% (3.0%) | 0 | 4.3% (1.7) |
|  | 70+ | 0 | 0 | 0 |
|  | Total | 3.7% (1.1) | 1.8%(0.91) | 2.9% (0.7) |
| Do you have shortness of breath when hurrying on the level or walking up a slight hill?) | 40-49 | 10.5% (3.0) | 17.8%(3.6) | 13.8% (2.3) |
|  | 50-59 | 6.1% (2.2) | 12.3%(3.7) | 9.04% (2.1) |
|  | 60-69 | 7.5% (3.2) | 6.7%(4.6) | 7.2% (2.7) |
|  | 70+ | 10.7% (4.6) | 5.5%(5.4) | 8.3% (3.5) |
|  | Total | 8.9% (1.7) | 13.2%(2.2) | 10.9% (1.3) |
| Any respiratory symptom (Any of cough, sputum, wheeze without cold, exertional breathlessness as above | 40-49 | 25.8% (4.2) | 25.3%(4.1) | 25.6% (2.9) |
|  | 50-59 | 21.1% (3.7) | 19.2%(4.4) | 20.2% (2.8) |
|  | 60-69 | 26.5% (5.2) | 22.7%(7.5) | 24.8% (4.4) |
|  | 70+ | 23.7% (6.7) | 18.4%(9.8) | 21.8% (5.8) |
|  | Total | 24.5% (2.4) | 22.4%(2.7) | 23.5%(1.9) |
| Functional limitation (Have breathing problems interfered with your usual daily activities) | 40-49 | 0.9% (0.9) | 5%(2.0) | 2.8% (1.1) |
|  | 50-59 | 0 | 3.7% (2.1) | 1.6% (0.9) |
|  | 60-69 | 1.3% (1.3) | 0 | 0.7% (0.7) . |
|  | 70+ | 1.8% (1.7) | 0 | 0.9% (0.9) |
|  | Total | 0.8% (0.5) | 3.4%(1.1) | 1.9%(0.5) |
| SE Standard Error | | | | |

**Table S2**: Age and gender stratified prevalence estimates for abnormal spirometry, among participant with full spirometry data

| Spirometric definition (Reference range) | Age group (n=516) | Male (n= 306)  Prevalence (SE) | Female (n=210)  Prevalence (SE) | Prevalence (SE) Total (n= 516) |
| --- | --- | --- | --- | --- |
| Estimated Population Prevalence of CAO defined using NHANES III (Post-BD FEV1/FVC < LLN) | 40-49 | 6.8(2.7) | 4.9(2.1) | 5.9(1.7) |
|  | 50-59 | 10.4(2.9) | 13.1(4.4) | 11.5(2.5) |
|  | 60-69 | 17.9(4.7) | 19.0(7.6) | 18.4(4.2) |
|  | 70+ | 5.6(3.9) | 21.1(10.8) | 13.4(6.0) |
|  | Total | 9.5(1.7) | 11.2(2.4) | 10.3(1.4) |
| Estimated Population Prevalence of CAO defined using locally derived reference range (Post-BD FEV1/FVC < LLN) | 40-49 | 4.7(2.3) | 3.0(1.7 | 3.9(1.5) |
|  | 50-59 | 4.3(1.9) | 3.1(2.2) | 3.8(1.4) |
|  | 60-69 | 11.9(3.9) | 15.4(7.1) | 13.4(3.8) |
|  | 70+ | - | 13.2(8.8) | 6.7(4.6) |
|  | Total | 5.2(1.3) | 6.2(1.9) | 5.7(1.1) |
| Estimated Population Prevalence of CAO Stage 2 or higher ((Post-BD FEV1/FVC < LLN and post-BD FEV1 < 80% predicted) (NHANES ref range) | 40-49 | 6.8(2.6) | 4.9(2.1) | 5.9(1.7) |
|  | 50-59 | 8.6(2.6) | 13.1(4.3) | 10.5(2.4) |
|  | 60-69 | 16.4(4.5) | 18.9(7.6) | 17.6(4.2) |
|  | 70+ | 5.6(3.9) | 13.2(8.8) | .9.4(4.9) |
|  | Total | 8.8(1.7) | 10.1(2.2) | 9.4(1.4) |
| Estimated Population Prevalence of CAO Stage 2 or higher COPD(Post-BD FEV1/FVC < LLN and post-BD FEV1 < 80% predicted) (locally derived ref range) | 40-49 | 1.1(1.1) | 3.0(1.7) | 2.0(1.0) |
|  | 50-59 | 3.5(1.7) | 1.8(1.8) | 2.8(1.3) |
|  | 60-69 | 8.8(3.4) | 3.9(3.9) | 6.7(2.6) |
|  | 70+ | - | 5.3(5.2) | 2.7(2.6) |
|  | Total | 2.9(0.9) | 3.1(1.3) | 3.0(0.8) |
| Estimated prevalence of spirometric restriction among unobustructed group (fev1fvcp_post>llnfev1fvcp_post_nhanes and fvc_post<llnfvc_nhanes) | 40-49 | 81.7(4.2) | 68.8(4.6) | 75.6(3.1) |
|  | 50-59 | 62.2(4.1) | 60.6(6.2) | 61.4(3.7) |
|  | 60-69 | 44.7(6.1) | 48.0(9.6) | 46.1(5.4) |
|  | 70+ | 36.4(7.9) | 39.4(12.6) | 38.1(7.5) |
|  | Total | 65.2(2.8) | 59.8(3.5) | 62.7(2.2) |
| Airway reversibility FEV1 increase ≥200ml AND ≥12% following bronchodilator | 40-49 | 1.3% (1.3) | 4.9% (2.2) | 3% (1.2) |
|  | 50-59 | 7.2% (2.5) | 9.1% (3.6) | 8.1% (2.1) |
|  | 60-69 | 5.9% (2.9) | 18.3% (7.4) | 11.2% (3.6) |
|  | 70+ | 7.5% (4.3) | 5.3% (5.2) | 6.4% (3.4) |
|  | Total | 4.4% (1.1) | 8% (1.9) | 6.1% (1.1) |

**Table S3:** univariable associations between respiratory symptoms and risk factors

| Variable | Usual Cough (n=66/ 596) | | Usual Sputum (n=67/ 596) | | Exertional dyspnoea (n=62/545) | | Wheeze without cold (n= 18/596) | |
| --- | --- | --- | --- | --- | --- | --- | --- | --- |
|  | Odds Ratio | 95% CI | Odds Ratio | 95% CI | Odds Ratio | 95% CI | Odds Ratio | 95% CI |
| Age Group |  |  |  |  |  |  |  |  |
| 40-49 | 1.0 | - | 1.0 | - | 1.0 | - | 1.0 | - |
| 50-59 | 1.15 | 0.61-2.16 | 0.60^±^ | 0.32-1.12 | 0.62 ^±^ | 0.33-1.17 | 0.35^±^ | 0.09-1.31 |
| 60-69 | 1.27 | 0.60- 2.70 | 0.82 | 0.39-1.72 | 0.48^±^ | 0.20- 1.19 | 1.10 | 0.37-3.18 |
| 70+ | 1.31 | 0.48-3.55 | 0.44^±^ | 0.16-1.21 | 0.57 | 0.21- 1.54 | - | - |
| Gender |  |  |  |  |  |  |  |  |
| Male | 1.0 | - | 1.0 | - | 1.0 | - | 1.0 | - |
| Female | 0.95 | 0.54- 1.65 | 0.61^±^ | 0.35-1.07 | 11.55^±^ | 0.89-2.71 | 0.48^±^ | 0.16- 1.41 |
| Year of education | 0.97^±^ | 0.92- 1.01 | 1.00 | 0.95- 1.05 | 1.05 ^*^ | 1.00-1.10 | 0.98 | 0.91-1.06 |
| Smoking |  |  |  |  |  |  |  |  |
| Never | 1.0 | - | 1.0 | - | 1.0 | - | 1.0 | - |
| Ever | 1.05 | .56- 1.99 | 2.38* | 1.36-4.15 | 1.45 | 0.76- 2.75 | 2.01^±^ | 0.70-5.78 |
| Smoking Status |  |  |  |  |  |  |  |  |
| Current | 1.57 | 0.67- 3.67 | 2.52* | 1.16-5.46 | 1.27 | 0.46-3.50 | 4.86 * | 1.56- 15.16 |
| Ex | 0.74 | 0.33- 1.69 | 2.28* | 1.16-4.48 | 1.57 | 0.74-3.33 | 0.33 | 0.04-2.61 |
| Packs per year | 1.0 | - | 1.0 | - | 1.0 | - | 1.0 | - |
| Never |  |  |  |  |  |  |  |  |
| <10 years | 0.86 | 0.33-2.17 | 2.36* | 1.15-4.76 | 1.36 | 0.63-2.94 | 0.78 | 0.10- 6.16 |
| > =10 years | 1.28 | 0.59-2.78 | 2.42* | 1.18-4.98 | 1.22 | 0.50- 3.01 | 3.48^*^ | 1.11-10.95 |
| BMI (kg/m^2^) |  |  |  |  |  |  |  |  |
| Underweight (BMI <18.5) | 0.92 | 0.21- 4.06 | 0.39 | 0.09-1.72 | 0.91 | 0.23-3.60 |  |  |
| Normal (18.5-25) | 1.0 | - | 1.0 | - | 1.0 | - | 1.0 | - |
| Overweight (25-30) | 1.21 | 0.63-2.32 | 0.73 | 0.39- 1.35 | 1.92 | 0.93-3.98 | 1.95 | 0.65- 5.82 |
| Obese (BMI>30) | 1.21 | 0.61- 2.42 | 0.58^±^ | 0.28-1.20 | 2.85* | 1.38-5.90 | 0.50 | 0.09-2.70 |
| Self-reported TB |  |  |  |  |  |  |  |  |
| No | 1.0 | - | 1.0 | - | 1.0 | - | - | - |
| Yes | 1.96 | 0.21-18.19 | 1.81 | 0.20-16.84 | 1.86 | 0.20 17.26 | - | - |
| Reported Hypertension |  |  |  |  |  |  |  |  |
| No | 1.0 | - | 1.0 | - | 1.0 | - | 1.0 | - |
| Yes | 0.73 | 0.35- 1.52 | 0.98 | 0.50- 1.94 | 0.97 | 0.49-1.93 | 1.30 | 0.41-4.19 |
| Reported Diabetes |  |  |  |  |  |  |  |  |
| No | 1.0 | - | 1.0 | - | 1.0 | - | 1.0 | - |
| Yes | 1.01 | 0.40- 2.57 | 2.88 * | 1.41-5.90 | 0.36^±^ | 0.08-1.55 | 1.45 | 0.40-5.28 |
| Reported Heart disease |  |  |  |  |  |  |  |  |
| No | 1.0 | - |  |  |  |  |  |  |
| Yes | 1.33 | 0.28- 6.30 | 0.53 | 0.07-4.26 | 1.03 | 0.13- 8.30 | - | - |
| Does household own home |  |  |  |  |  |  |  |  |
| No | 1.0 | - | 1.0 | - | 1.0 | - | 1.0 | - |
| Yes | 1.38 | 0.77-2.47 | 1.01 | 0.56- 1.81 | 1.09 | 0.60- 1.99 | 0.63 | 0.19-2.03 |
| Access to own water supply |  |  |  |  |  |  |  |  |
| No | 1.0 | - | 1.0 | - | 1.0 | - | 1.0 | - |
| Yes | 0.89 | 0.49-1.62 | 0.68 | 0.37-1.23 | 0.26* | 0.11-0.59 | 0.78 | 0.28-2.23 |
| Does Household have a flush Toilet |  |  |  |  |  |  |  |  |
| No | 1.0 | - | 1.0 | - | 1.0 | - | 1.0 | - |
| Yes | 0.84 | 0.44-1.58 | 1.63^±^ | 0.79- 3.36 | 0.58^±^ | 0.32-1.06 | 7.30^±^ | 0.95-56.07 |
| Number of people living in house | 1.06^±^ | 0.99-1.13 | 1.03 | 0.97- 1.10 | 1.02 | 0.93-1.13 | 0.96 | 0.83-1.11 |
| Biomass exposure |  |  |  |  |  |  |  |  |
| No | 1.0 | - | 1.0 | - | 1.0 | - |  |  |
| Yes | 1.90^±^ | 0.80-4.51 | 1.78 | 0.67-4.68 | 3.51* | 1.19- 10.38 | 1.46 | 0.30- 7.07 |
| Working in a Farm |  |  |  |  |  |  |  |  |
| No | 1.0 | - | 1.0 | - | 1.0 | - |  |  |
| Yes | 0.97 | 0.50- 1.90 | 1.30 | 0.70- 2.41 | 0.55^±^ | 0.25- 1.22 | 1.22 | 0.36- 4.16 |
| *indicates a P<0.05 ^±^ indicates a P<0.2 | | | | | | | | |

**Table S4** Multivariable associations between respiratory symptoms and risk factors, all variables significant at level <0.2 are included

| Variable | Usual Cough (n=66/ 596) | | Usual Sputum (n=67/ 596) | | Exertional dyspnoea (n=62/545) | | Wheeze without cold (n= 18/596) | |
| --- | --- | --- | --- | --- | --- | --- | --- | --- |
|  | Odds Ratio | 95% CI | Odds Ratio | 95% CI | Odds Ratio | 95% CI | Odds Ratio | 95% CI |
| Age Group |  |  |  |  |  |  |  |  |
| 40-49 | 1.0 | - | 1.0 | - | 1.0 | - | 1.0 | - |
| 50-59 | 1.05 | 0.54-2.03 | 0.36* | 0.17- 0.75 | 0.49 | 0.22-1.09 | 0.34 | 0.09- 1.33 |
| 60-69 | 0.99 | 0.43- 2.29 | 0.39* | 0.16-0.94 | 0.81 | 0.31-2.14 | 1.28 | 0.45- 3.70 |
| 70+ | 0.97 | 0.36- 2.66 | 0.41 | 0.15- 1.11 | 0.29 | 0.05-1.72 | - | - |
| Gender |  |  |  |  |  |  |  |  |
| Male | 1.0 | - | 1.0 | - | 1.0 | - | 1.0 | - |
| Female | 0.79 | 0.43- 1.45 | 0.93 | 0.44- 1.96 | 1.37 | 0.63-2.99 | 0.61 | 0.19-1.98 |
| Year of education | 0.96 | 0.91- 1.02 |  |  | 1.03 | 0.97- 1.09 |  |  |
| Smoking Status |  |  |  |  |  |  |  |  |
| Never | - | - | 1.0 | - | 1.0 | - | 1.0 | - |
| Current | - | - | 2.25 | 0.77- 6.55 |  |  | 3.49* | 1.02-11.96 |
| Ex-Smoker | - | - | 2.67* | 1.09- 6.50 |  |  | 0.26 | 0.03-2.07 |
| BMI (kg/m^2^) |  |  |  |  |  |  |  |  |
| Underweight (BMI <18.5) | - | - | 0.19 | 0.03- 1.43 | 0.53 | 0.10-2.74 | - | - |
| Normal (18.5-25) | - | - | 1.0 | - | 1.0 | - | - | - |
| Overweight (25-30) | - | - | 0.64 | 0.30- 1.35 | 2.09 | 0.91-4.76 | - | - |
| Obese (BMI>30) | - | - | 0.75 | 0.34- 1.68 | 2.31 | 0.94- 5.66 | - | - |
| Reported Diabetes |  |  |  |  |  |  |  |  |
| No | - | - | 1.0 | - | 1.0 | - |  |  |
| Yes | - | - | 4.04* | 1.82- 8.96 | 0.26 | 0.03-1.94 |  |  |
| Does Household have a flush Toilet |  |  |  |  |  |  |  |  |
| No | - | - | 1.0 | - | 1.0 | - | 1.0 | - |
| Yes | - | - | 1.24 | 0.53- 2.87 | 0.62 | 0.29-1.33 | 5.61 | 0.71-44.36 |
| Number of people living in house | 1.05 | 0.99- 1.12 | - | - | - | - | - | - |
| Any biomass exposure |  |  |  |  |  |  |  |  |
| No | 1.0 | - | 1.0 | - | 1.0 | - | - | - |
| Yes | 1.69 | 0.70- 4.09 | 1.93 | 0.71- 5.23 | 4.56 * | 1.44- 14.43 | - | - |
| Working in a Farm |  |  |  |  |  |  |  |  |
| No | - | - | - | - | 1.0 | - | - | - |
| Yes | - | - | - | - | 0.90 | 0.36- 2.28 | - | - |
| *indicates a P<0.05 | | | | | | | | |

**Table S5** Univariable and multivariable associations of risk factors with CAO defined using NHANES III (Post-BD FEV1/FVC < LLN ; n=53/516)

| Variable | Univariable association | | Multivariable association | |
| --- | --- | --- | --- | --- |
|  | Odds Ratio | 95% CI | Odds Ratio | 95% CI |
| Age Group |  |  |  |  |
| 40-49 | 1.0 | - | 1.0 | - |
| 50-59 | 2.08 ^±^ | 0.95- 4.53 | 2.13 | 0.84 - 5.41 |
| 60-69 | 3.58 * | 1.56- 8.22 | 3.16 * | 1.20 - 8.32 |
| 70+ | 2.47 | 0.76- 9.10 | 1.91 | 0.60- 6.10 |
| Gender |  |  |  |  |
| Male | 1.0 | - | 1.0 | - |
| Female | 1.20 | 0.65- 2.23 | 1.31 | 0.61- 2.84 |
| Level of education |  |  |  |  |
| None | 1.0 | - | 1.0 | - |
| Primary school | 0.46^±^ | 0.21- 1.021 | 0.61 | 0.27- 1.34 |
| Middle school | 0.72 | 0.27- 1.91 | 1.23 | 0.40- 3.81 |
| High school or above | 0.32* | 0.134- 0.76 | 0.71 | 0.28- 1.78 |
| Self-reported TB |  |  |  |  |
| No | 1.0 | - | - | - |
| Yes | 4.533718^±^ | 0.72- 28.68 | 0.08 | 0.01- 0.59 |
| Reported Hypertension |  |  |  |  |
| No | 1.0 | - | - | - |
| Yes | 0.75 | 0.35- 1.61 | - | - |
| BMI (kg/m2) |  |  |  |  |
| Underweight (BMI<18.5) | 2.04^±^ | 0.72- 5.77 | 1.87 | 0.66- 5.30 |
| Normal (BMI 18-25) |  |  | 1.0 | - |
| Overweight (BMI 25-30) | 0.38* | 0.18- 0.82 | 0.43 | 0.18-1.01 |
| Obese (BMI >30) | 0.34* | 0.13- 0.89 | 0.35 | 0.11- 1.17 |
| Smoking Status |  |  |  |  |
| Never | 1.0 | - | - | - |
| Ever | 0.94 | 0.46- 1.92 | - | - |
| Smoking Status |  |  |  |  |
| Never | 1.0 | - | 1.0 | - |
| Current | 1.21 | 0.48- 3.05 | 1.95 | 0.83- 4.59 |
| Ex | 0.76 | 0.29- 1.97 | 0.72 | 0.32- 1.60 |
| Smoking packs years |  |  |  |  |
| <10 Years |  |  |  |  |
| <= 10 Years | 0.51^±^ | 0.19- 1.36 | - | - |
| Home ownership | 1.0 | - | - | - |
| No | 1.0 | - | - | - |
| Yes | 1.11 | 0.55- 2.23 | - | - |
| Access to private water supply (indoors or outdoors) |  |  |  |  |
| No | 1.0 | - | - | - |
| Yes | 1.60 ^±^ | 0.41- 1.82 | - | - |
| Household has flush toilet |  |  |  |  |
| No | 1.0 | - | - | - |
| Yes | 0.86 | 0.41- 1.82 | - | - |
| Number of people living in house | 1.051 | 0.97- 1.14 | - | - |
| Any biomass exposure |  |  |  |  |
| No |  |  | - | - |
| Yes | 1.80 | 0.65- 4.97 | - | - |
| Use of firewood in cooking >6 month |  |  |  |  |
| No | 1.0 | - | - | - |
| Yes | 0.60^±^ | 0.30-0.17 | - | - |
| *^±^ indicates P<0.2,*p<0.05. CI, confidence interval; OR, odds ratio; FEV_1_, forced expiratory volume in 1 second; FVC, forced vital capacity ratio; LLN, lower limit of normal* | | | | |

**Table S6** Univariable and multivariable associations of risk factors with Stage 2 or higher CAO defined using NHANES III (Post-BD FEV1/FVC < LLN and post-BD FEV1 < 80% predicted; n=49/516)

| Variable | Univariable association | | Multivariable association | |
| --- | --- | --- | --- | --- |
|  | Odds Ratio | 95% CI | Odds Ratio | 95% CI |
| Age Group |  |  |  |  |
| 40-49 | 1.0 | - | 1.0 | - |
| 50-59 | 2.27* | 1.06- 4.87 | 1.86 | 0.73- 4.77 |
| 60-69 | 5.82* | 2.68-12.67 | 2.78* | 1.07- 7.26 |
| 70+ | 3.13* | 1.13- 8.65 | 0.91 | 0.21-3.92 |
| Gender |  |  |  |  |
| Male | 1.0 | - | 1.0 | - |
| Female | 0.76 | 0.43-1.34 | 1.67 | 0.76- 3.66 |
| Level of education |  |  | 1.0 | - |
| None | 1.0 | - | 0.62 | 0.27- 1.44 |
| Primary school | 0.53^±^ | 0.24-1.17 | 1.03 | 0.32-3.36 |
| Middle school | 0.77 | 0.28-2.12 | 0.69 | 0.26- 1.86 |
| High school or above | 0.35* | 0.14-0.85 | 1.0 | - |
| Self-reported TB |  |  |  |  |
| No | 1.0 | - | 1.0 | 0 |
| Yes | 3.58^±^ | 0.57- 22.51 | 0.07* | 0.01- 0.48 |
| BMI (kg/m2) |  |  |  |  |
| Underweight (BMI<18.5) | 1.52 | 0.57- 4.03 | 1.84 | 0.64- 5.27 |
| Normal (BMI 18-25) | 1.0 | - | 1.0 | - |
| Overweight (BMI 25-30) | 0.43* | 0.22-0.83 | 0.48 * | 0.23- 0.10 |
| Obese (BMI >30) | 0.29* | 0.12- 0.71 | 0.43 | 0.16- 1.20 |
| Smoking |  |  |  |  |
| Never | 1.0 | - |  |  |
| Ever | 1.26 | 0.69- 2.31 |  |  |
| Smoking Status |  |  |  |  |
| Never | 1.0 | - | 1.0 | - |
| Current | 1.96^±^ | 0.91- 4.22 | 2.28 | 0.96-5.41 |
| Ex | 0.82 | 0.35-1.90 | 0.59 | 0.23-1.49 |
| Household has flush toilet |  |  |  |  |
| No | 1.0 | - | - | - |
| Yes | 0.99 | 0.52- 1.87 | - | - |
| Number of people living in house | 1.06 ^±^ | 0.98-1.13 | 1.05 | 0.96-1.16 |
| Any biomass exposure |  |  |  |  |
| No | 1.0 | - | - | - |
| Yes | 1.69 | 0.66-4.31 | - | - |
| Working in Farming |  |  |  |  |
| No | 1.0 | - |  |  |
| Yes | 1.53^±^ | 0.83-2.80 | 0.61 | 0.32-1.16 |
| Wealth score/Mokken scale | 0.94 | 0.83-1.06 | 0.96 | 0.83-1.11 |
| *^±^ indicates P<0.2,*p<0.05. CI, confidence interval; OR, odds ratio; FEV_1_, forced expiratory volume in 1 second; FVC, forced vital capacity ratio; LLN, lower limit of normal* | | | | |
