## Supplementary figures and images for "Prevalence and determinants of chronic respiratory diseases in adults in Khartoum State, Sudan"

### Figure 1

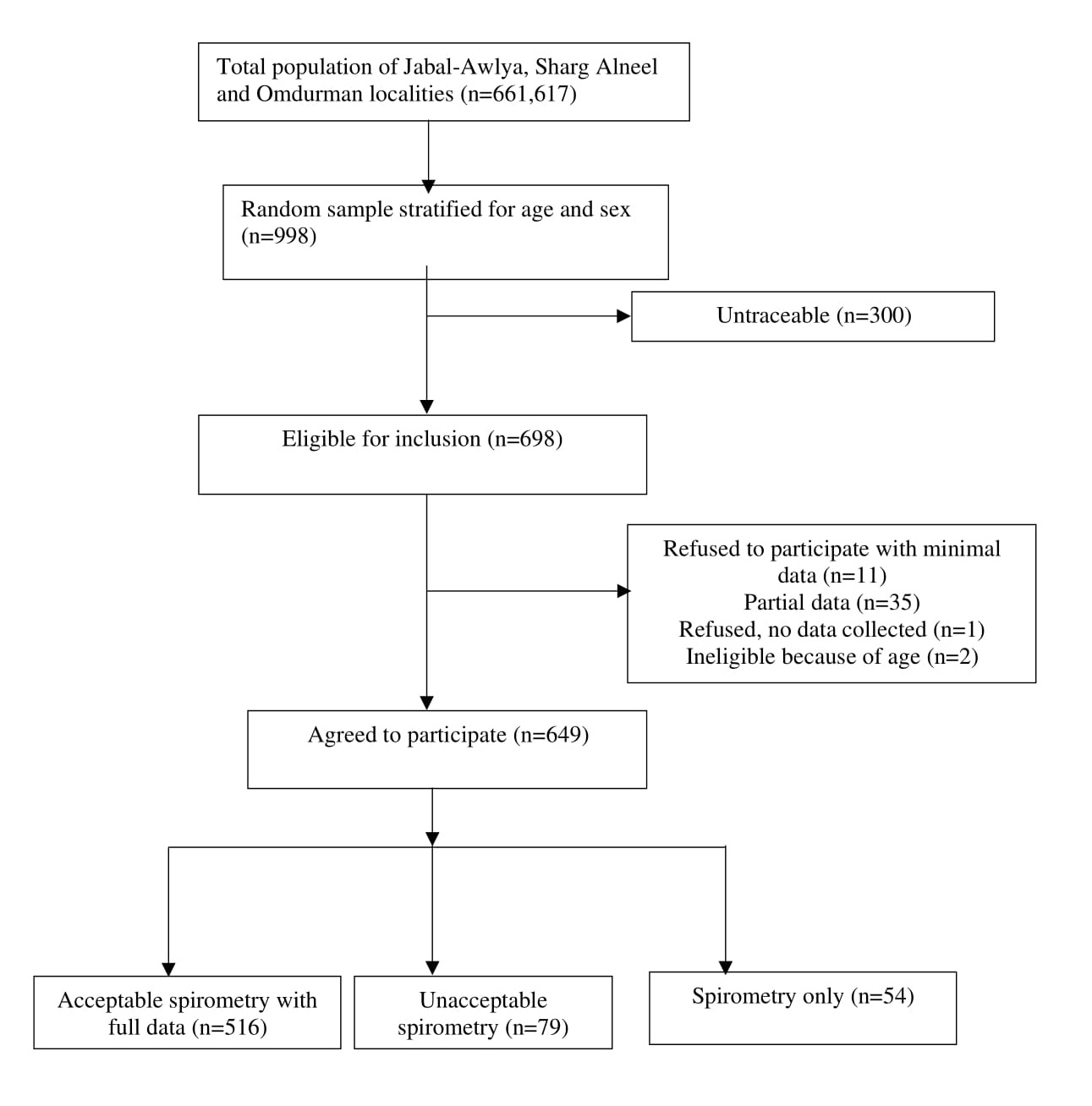

### Figure 2

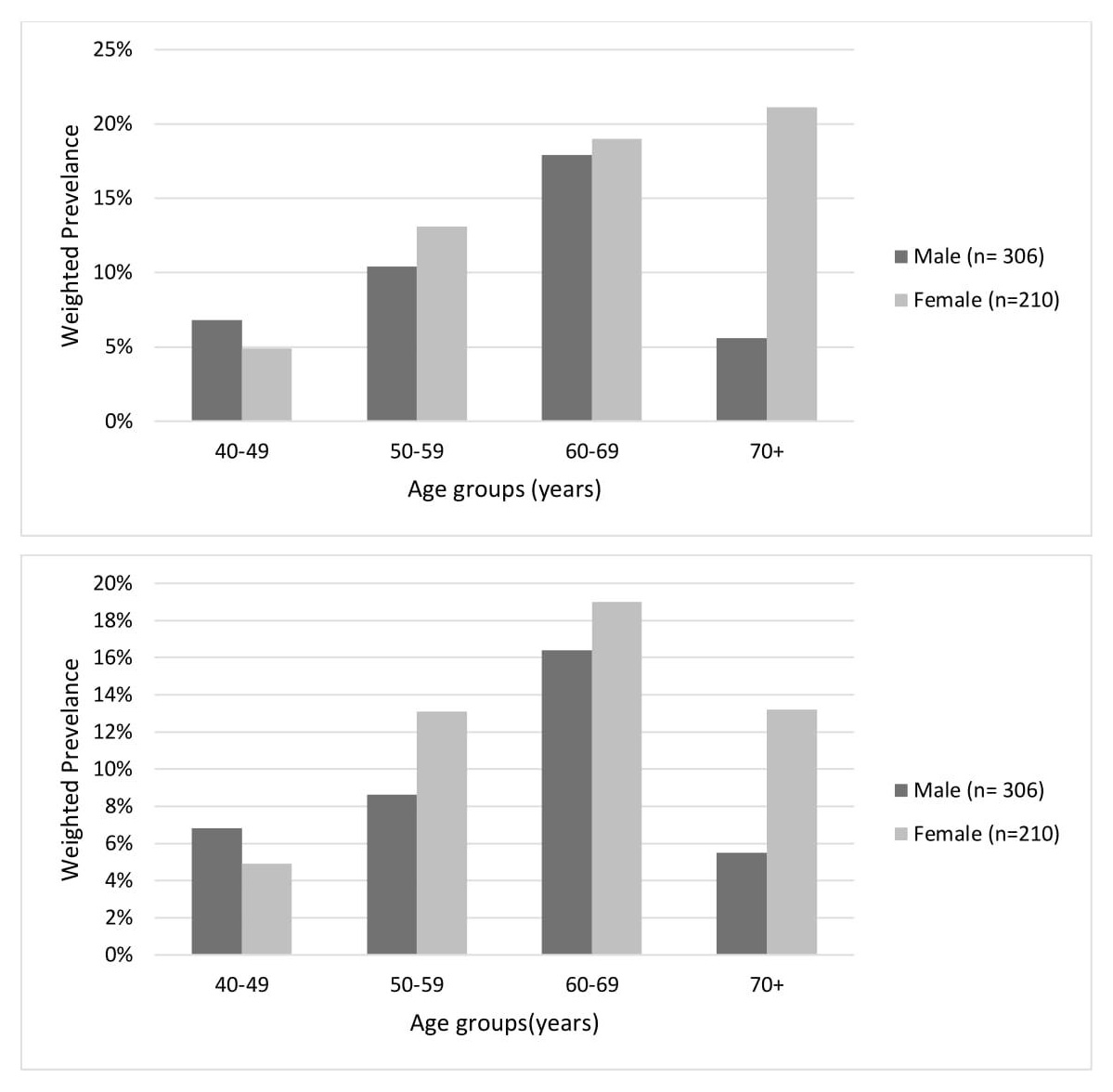
